# The effects of modifiable maternal pregnancy exposures on offspring molar-incisor hypomineralisation: A negative control study

**DOI:** 10.1101/2021.10.27.21265567

**Authors:** Qui-Yi Lim, Kurt Taylor, Tom Dudding

## Abstract

**Objectives:**

1. Explore associations between modifiable maternal pregnancy exposures: pre-pregnancy body mass index (BMI), pregnancy smoking and alcohol consumption with offspring molar-incisor hypomineralisation (MIH).
2. Use negative control analyses to explore for the presence of confounding.

**Methods:** Using prospectively collected data from Avon Longitudinal Study of Parents and Children (ALSPAC), we performed logistic regression to explore confounder adjusted associations between maternal pre-pregnancy BMI and smoking and alcohol consumption during pregnancy with MIH. We compared these with negative control exposure (paternal BMI, smoking and alcohol) and outcome (offspring dental trauma) analyses.

**Results:** 5,536 mother/offspring pairs were included (297 MIH cases [5.4%]). We found a weak, positive association between maternal mean BMI and offspring MIH (OR per 1□kg/m^2^ difference in BMI: 1.04, 95%CI: 1.00, 1.08). Results of subsequent analyses suggested this effect was non-linear and being driven by women in the highest BMI quintile (OR for women in the highest BMI quintile versus the lowest: 1.61 95%CI: 1.02, 2.60). Negative control analyses showed no evidence of an association between paternal BMI and offspring MIH (OR: 0.94, 95%CI: 0.89,1.00) and maternal BMI and offspring dental trauma (OR: 0.99, 95%CI: 0.96, 1.02). There was no clear evidence of an association for maternal smoking (OR: 0.76, 95%CI: 0.46,1.22) or alcohol consumption (OR: 0.79, 95%CI: 0.56, 1.21) with offspring MIH with results imprecisely estimated.

**Conclusion:** We found evidence of a possible intrauterine effect for high maternal pre-pregnancy BMI on offspring MIH, but no robust evidence of an intrauterine effect for maternal pregnancy smoking or alcohol consumption. A key limitation includes possible misclassification of MIH. Replication of these results is warranted.

## 1. Introduction

Molar-incisor hypomineralisation (MIH) is a developmental, qualitative enamel defect caused by reduced mineralisation and inorganic enamel components. It primarily affects one or more first permanent molars (FPMs), and frequently involves incisors (Weerheijm, 2003). MIH is often diagnosed around 6-7 years of age when the FPMs and central incisors erupt into the oral cavity. Due to the poor quality enamel, affected teeth present aesthetically concerning with discoloured creamy-white or yellow-brown demarcations (Weerheijm, 2003). These teeth also carry pathological concerns as they are often hypersensitive, susceptible to post-eruptive breakdown (Bullio Fragelli et al., 2015) and rapid caries progression (Americano et al., 2017). Consequently, MIH-affected teeth often have poor prognosis and are frequently extracted before adulthood.

The reported prevalence of MIH varies between 2.4 and 40.2%, with a global mean of 13.1% (95% confidence interval (CI): 11.8, 14.5) (Schwendicke et al., 2018). The literature remains inconclusive around the aetiology of MIH; however, it is generally accepted that causes are likely to be multifactorial. Many studies propose a genetic predisposition (Jeremias et al., 2016). Although, the clinical presentation of these localised, asymmetrical lesions indicate a further cause of systemic or environmental origin which disrupts enamel formation. The development of FPMs begins *in utero* and continues to develop 2-3 years after birth (Schuurs, 2012; Welbury et al., 2018). With this prolonged developmental time window, a range of prenatal, perinatal, and post-natal factors have been investigated. These include maternal illness, low offspring birth weight, childhood antibiotic exposure, and many more (Fatturi et al., 2019; Silva et al., 2016). Despite FPMs developing in utero there is little research on maternal exposures during pregnancy.

To our knowledge, there have been no studies that have investigated the effect of maternal body mass index (BMI) on MIH. Maternal pre-pregnancy obesity is known to affect foetal development (Leddy et al., 2008; Maffeis & Morandi, 2017). FPMs develop *in utero*, therefore, it is plausible that changes in BMI may influence dental development. Effects of pregnancy smoking and alcohol consumption have been well established with birth defects associated with craniofacial and dental abnormalities, such as oral facial clefts for smoking (Little et al., 2004), and foetal alcohol syndrome for alcohol (Sant’Anna & Tosello, 2006). Effects of pregnancy smoking and alcohol on MIH have also been investigated. However, recent systematic reviews on these risk factors concluded that there is no significant evidence to show an association with MIH (Fatturi et al., 2019; Silva et al., 2016). Conversely, the primary literature exists to be conflicting and the strength of evidence is limited. Previous studies have been limited due to a lack of detail and consistency in the maternal exposures and MIH outcomes investigated (Silva et al., 2016) which makes comparisons between studies difficult. Many studies have also failed to adjust for confounding variables, have had limited statistical power, and predominantly use retrospective study designs which are prone to common epidemiological biases, such as recall bias. Therefore, we cannot determine whether these reflect the magnitude of the causal effect, or if these results are biased by residual confounding, systematic reporting bias or measurement bias.

Negative controls are used in epidemiology to detect potential confounding after adjusting for measured confounders (Lipsitch et al., 2010). There are two types of negative control experiments: a negative control exposure, shown in **Figure 1A**, and a negative control outcome, shown in **Figure 1B** (Lipsitch et al., 2010). The idea of these negative control experiments is to substitute a condition with an exposure or outcome variable that (i) shares similar confounding structures as the ‘real study’ (the original association), and (ii) does not have a plausible biological link with the association of interest (Gage et al., 2016). Under these two assumptions, it can be assumed that the ‘real study’ and negative control experiment are perfectly comparable, and we can therefore expect results of the negative controls to produce a weaker or no association if there were to be a true causal effect (Lipsitch et al., 2010). Considering these two criteria, we used paternal exposures measured around the time of pregnancy as a negative exposure control (**Figure 1A**), and offspring dental trauma as a negative control outcome (**Figure 1B**). Parental negative exposure controls have been used previously to explore the intrauterine effects of maternal exposures and offspring outcomes (Brand et al., 2019; K. Taylor et al., 2021).

**Figure 1:**
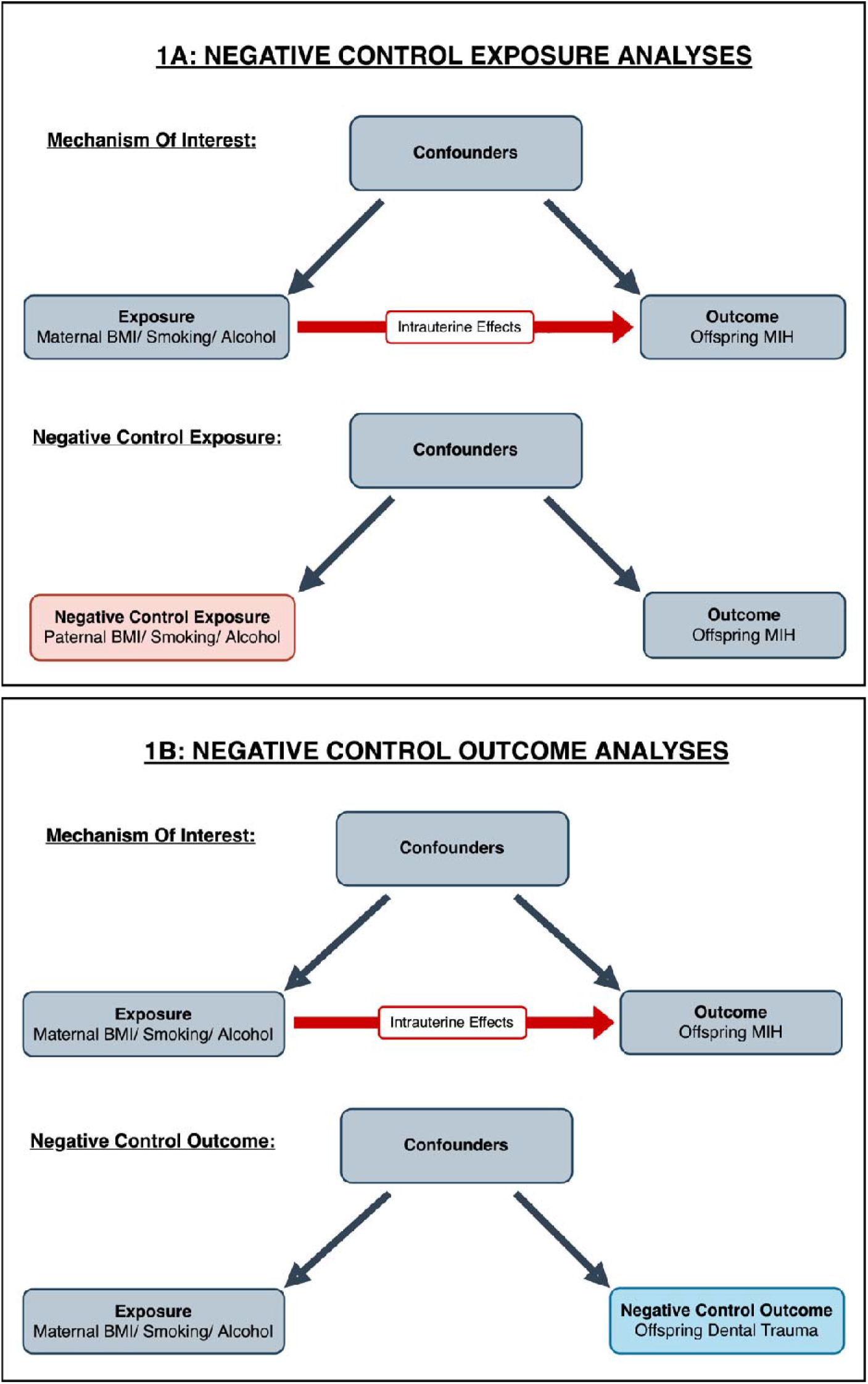
Diagrams to show negative control experiments. **1A** shows how a negative control exposure, paternal BMI/ smoking/ alcohol can be used to investigate effects of intrauterine exposures on MIH birth outcomes. The paternal exposure has the same incoming arrows as the maternal exposure of interest, but has no arrow to the offspring MIH outcome – this means that any association observed between the paternal exposure and MIH outcome of interest will be due to the confounding variables in the model. The assumptions of this approach are that: (i) measured and unmeasured confounders influence the exposures in the same direction and with a similar magnitude in mothers and fathers and (ii) there is no plausible reason why the exposure in the father would affect the offspring outcome (or at a minimum the paternal association would be much weaker than in the mother) (**Figure 1A**). **1B** shows how a negative control outcome, offspring dental trauma can be used to investigate effects of intrauterine exposures on MIH birth outcomes. Offspring dental trauma has the same incoming arrows as the MIH outcome of interest, but has no arrow from the maternal exposure of interest – this means that any association observed between the maternal exposure and offspring dental trauma will be due to the confounding variables in the model. Each exposure variable (maternal and paternal BMI, smoking and alcohol consumption) and outcome variable (offspring MIH and dental trauma) will be looked at separately. We used offspring dental trauma as a negative outcome control under the assumptions that: (i) offspring dental trauma shares similar confounding structures to offspring MIH and (ii) maternal exposures during pregnancy could not plausibly cause offspring dental trauma.

In summary, the exact causes of MIH are unclear and intrauterine factors via maternal pregnancy exposures could be relevant. There is a need for more prospective research using study designs that are reliably able to assess the presence of confounding. Identifying modifiable risk factors for MIH is important for improving aetiological understanding and developing preventive interventions to reduce disease burden.

The aims of this study were to:

1. Explore associations between maternal pre-pregnancy BMI, pregnancy smoking and alcohol consumption with offspring MIH.
2. Use negative control exposure and outcome analyses to explore for the presence of residual confounding.

## 2. Methods

### 2.1 Data

We used data from Avon Longitudinal Study of Parents and Children (ALSPAC), an ongoing, multigenerational, prospective birth cohort study. 14, 541 pregnant women residing in Avon, UK with expected delivery dates between 1^st^ April 1991 and 31^st^ December 1992 were recruited. Two further rounds of recruitment totalled 15, 454 pregnant women and 14, 901 offspring alive at 1 year of age were enrolled in the study (**Figure 2**) (Boyd et al., 2013; Fraser et al., 2013). Participants have been regularly followed up through questionnaires and clinic visits at the time of pregnancy in the mothers and fathers, and subsequent offspring from early life through to adulthood. The study website contains details of all the data that is available through a fully searchable data dictionary and variable search tool (http://www.bristol.ac.uk/alspac/researchers/our-data/). Ethical approval for the study was obtained from the ALSPAC Ethics and Law Committee and Local Research Ethics Committees.

**Figure 2:**
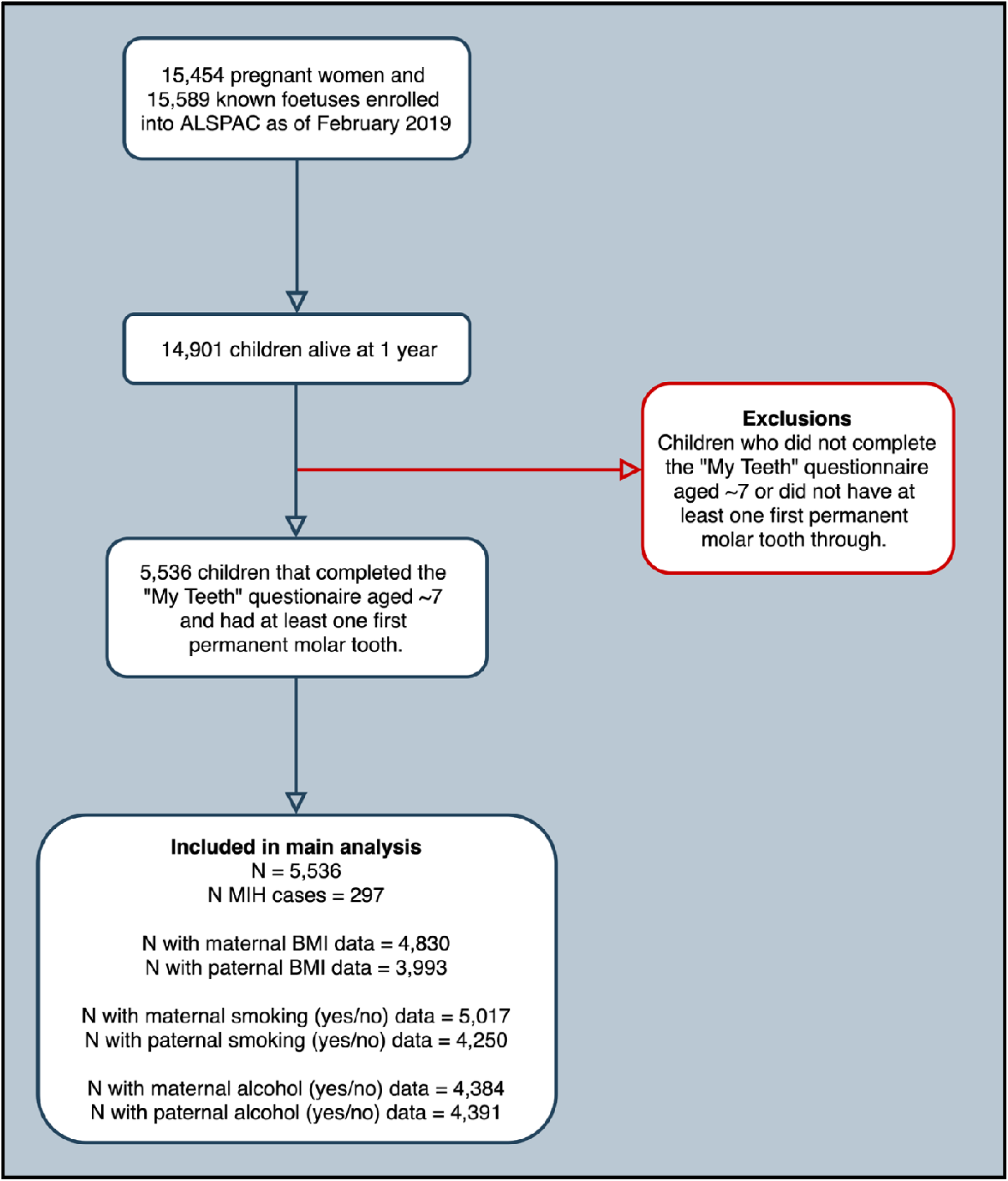
Study flow diagram illustrating participant selection in the ALSPAC cohort.

### 2.2 Participant inclusion criteria

At age 7 and 10, participants completed self-report questionnaires on dental health, with assistance from their parents and use of a mirror (**Figure S1, S2**). A subset of participants who had: (i) completed the questionnaire and (ii) had at least one first permanent molar tooth through by age 7 were included in this study. In total, 5,536 mother-offspring pairs were included. The flow of participants through the study are illustrated in **Figure 2**.

### 2.3 Exposures

#### Primary exposure: Maternal BMI, smoking and alcohol consumption

Maternal BMI was calculated using self-reported pre-pregnancy weight and height using data from the questionnaire completed around 12-weeks’ gestation. This was found to be correlated with the clinically measured pre-pregnancy weight and height. For analyses, BMI was used as a continuous variable. Maternal pregnancy smoking and alcohol consumption was assessed using questionnaires at 18- and 32-weeks’ gestation. Mothers reported the number of times smoked per day and how often they consumed alcoholic drinks, measured in glasses per week for each trimester. For smoking, data from each trimester was categorised into binary variables (yes/no), and used to generate one variable, categorised as “any pregnancy smoking” (yes/no). For alcohol consumption, data were available for the first and third trimester. These were used to generate one binary variable, “any pregnancy alcohol consumption” (yes/no). The rationale for including all three trimesters for these exposures is because the development of the FPMs begins 3.5-4 months *in utero* and continues after birth (Schuurs, 2012; Welbury et al., 2018). Therefore, it is plausible that the intrauterine environment throughout the entire pregnancy could influence offspring MIH.

#### Negative control exposures: Paternal BMI, smoking and alcohol consumption

We used paternal BMI, smoking and alcohol consumption during pregnancy as negative control exposures. All data used was self-reported by the partners at 18-weeks’ gestation. Paternal BMI was calculated using self-reported weight and height, and used as a continuous variable. Paternal smoking habits were measured as the number of times smoked at the start of pregnancy, and defined as a binary variable, “any partner smoking” (yes/no). Paternal alcohol consumption was measured using questions asking how often they consumed alcoholic drinks in the past 3 months, measured in glasses per week, and defined as a binary variable “any alcohol consumption” (yes/no).

### 2.4 Outcomes

#### Primary outcome: Offspring MIH

We defined MIH by using questions related to the child’s “6-year molars” from the age 7 and 10 self-report questionnaires **(Figure S1, S2)**. In the age 7 questionnaire, participants were asked to specifically check if each FPM had come through and identify if and any of these had come through brown (**Figure S1**). Using these questions, we defined cases of MIH as “any child who had at least one FPM that came through looking brown”. This was classified as a binary outcome, “any MIH” (yes/no).

#### Negative control outcome: Offspring dental trauma

We used offspring dental trauma as a negative control outcome. Questions related to “accidents to your teeth” from the questionnaire answered at age 10 were used (**Figure S2B**). Dental trauma was defined as any child whose teeth became “loose”, “chipped” or “knocked out” when they banged their top adult teeth. This negative control outcome was defined as binary, “any dental trauma” (yes/no).

### 2.5 Confounders

Analyses were adjusted for confounders which have previously shown to be a cause or plausible influence on maternal BMI, smoking and alcohol exposure and the offspring MIH outcome. The confounders adjusted for in the maternal and paternal analyses include age, education, parity (maternal), smoking (for BMI and alcohol analyses) and alcohol consumption (for BMI and smoking analyses). BMI, smoking, and alcohol were defined as above. Maternal and paternal age were kept as continuous variables. Parity was measured using data from the mother’s questionnaire, answered at 18-weeks’ gestation. We used questions regarding the mother’s previous pregnancies, which asked how many times the mother had previously been pregnant. This was used to define parity as nulliparous and multiparous. Educational attainment was used as a proxy measure of parents’ socioeconomic status. We used questions concerning educational qualifications for both the mothers, and their partners from the mother’s questionnaire completed around 32-weeks’ gestation. Educational attainment was classified into three categories, low (none/ CSE), medium (vocational/ O/ A level) and high (university degree).

### 2.6 Statistical Analysis

Analyses were conducted using R (Version 4.0.3). Analysis steps were discussed and agreed upon prior to any analyses taken place and documented in a pre-specified analysis plan (Kurt Taylor et al., 2021). We used logistic regression with maximal numbers (i.e. numbers included in each model are likely to vary due to missing data on exposure/ outcome or confounders). All analyses were run unadjusted (model 1), confounder adjusted (model 2), and confounder adjusted with the addition of the other parent’s exposure (BMI/ smoking/ alcohol consumption) (model 3). Model 3 produces a maternal association that adjusts for maternal confounders as well as the paternal exposure, and similarly a paternal association adjusting for paternal confounders and the maternal exposure, referred to as ‘other parent adjusted’. The reason for mutual parent adjustment is that parental BMI, smoking and alcohol consumption may associate with one another through assortative mating, shared social and environmental background, and modelling for each other’s behaviours (Sharp & Lawlor, 2019).

For the negative control exposure analyses, all three models of analyses were performed, then maternal and paternal exposures were directly compared with MIH outcomes by observing point estimates and 95% Confidence Intervals (CI)s. In the negative control outcome analyses, the first two models of analyses were performed, then maternal exposures were directly compared with MIH and dental trauma (negative control) outcomes.

To assess for deviation from linearity in the BMI-MIH association, BMI was split into quintiles for mothers and fathers. This BMI quintile variable was treated as both continuous and categorical in logistic regression models. We report p-values for a linear trend. (**Table S2**)

We performed a sensitivity analysis defining cases as those that had affected FPMs and permanent incisors using the age 10 questionnaire. Analyses were also performed on complete cases (cases with no missing data on exposure, outcome, or confounders) to investigate the influence of missing data (missing data reported in **Table S1**). Complete case outcomes were compared with the main analyses, which is reported in the supplementary data **(Table S3)**.

## 3. Results

### Participant characteristics

**Figure 2** shows selection of participants from the ALSPAC cohort into the study population. **Table 1** shows the distributions of offspring, maternal and paternal characteristics of this study population. 5,536 offspring had completed the dental questionnaires and had at least one FPM tooth. The prevalence of offspring MIH was 5.4% (**Table 1**), and 40 of the 297 (18.8%) MIH cases had both molars and incisors affected at age 10. The prevalence of dental trauma was 13.5%. Mean BMI was 22.3kg/m^2^ and 25.1kg/m^2^ for mothers and fathers, respectively. 21.4% of mothers smoked during pregnancy and 31.6% of fathers smoked around the time of pregnancy. 73.5% of mothers consumed any alcohol during pregnancy and 96.1% of fathers consumed any alcohol around the time of pregnancy.

**Table 1:** Participant characteristics.

|  | All | MIH | No MIH |
| --- | --- | --- | --- |
| **Offspring** |  |  |  |
| **Total N** | 5536 | 297 | 5239 |
| **N (%) MIH case**<br>(molars only) | 297 (5.4) | 297 (100.0) |  |
| **N (%) MIH sensitivity**<br>(molars and incisors) | 40 (0.7) | 40 (18.8) |  |
| **N (%) Dental trauma** | 621 (13.5) | 0 (0.0) | 621 (14.4) |
| **Maternal** |  |  |  |
| **Mean (SD) age (years)** | 29.5 (4.5) | 29.2 (4.6) | 29.53 (4.5) |
| **Mean (SD) BMI (kg/m<sup>2</sup>)** | 22.3 (4.1) | 22.9 (4.8) | 22.28 (4.0) |
| **N (%) Pregnancy Smoking** |  |  |  |
| Trimester 1 | 988 (18.2) | 67 (23.0) | 921 (18.0) |
| Trimester 2 | 782 (14.4) | 57 (19.6) | 725 (14.1) |
| Trimester 3 | 747 (15.1) | 50 (19.5) | 697 (14.9) |
| Any smoking | 1,072 (21.4) | 73 (27.3) | 999 (21.0) |
| **N (%) Pregnancy Alcohol** |  |  |  |
| Trimester 1 | 3,001 (55.6) | 153 (53.3) | 2,848 (55.7) |
| Trimester 3 | 1,074 (35.0) | 47 (29.7) | 1,027 (35.3) |
| Any alcohol | 3,224 (73.5) | 166 (70.9) | 3,058 (73.7) |
| **N (%) Parity** | 2,841 (53.0) | 152 (52.8) | 2,689 (53.0) |
| **Education** |  |  |  |
| Low: None/ CSE | 710 (13.3) | 55 (19.7) | 655 (12.9) |
| Medium: Vocational/ O/ A Level | 3,713 (69.7) | 184 (65.9) | 3,547 (69.9) |
| High: Degree | 910 (17.0) | 40 (14.3) | 870 (17.2) |
| **Paternal** |  |  |  |
| **Mean (SD) age (years)** | 31.6 (5.6) | 31.3 (5.9) | 31.57 (5.5) |
| **Mean (SD) BMI (kg/ m<sup>2</sup>)** | 25.1 (3.2) | 24.9 (3.2) | 25.13 (3.2) |
| **N (%) Pregnancy Smoking** |  |  |  |
| Any smoking | 1,344 (31.6) | 71 (33.8) | 1,273 (31.5) |
| **N (%) Pregnancy Alcohol** |  |  |  |
| Any alcohol | 4,219 (96.1) | 205 (95.3) | 4014 (96.1) |
| **N (%) Education** |  |  |  |
| Low: None/ CSE | 996 (19.1) | 65 (24.3) | 931 (18.8) |
| Medium: Vocational/ O/ A- level | 3,043 (58.2) | 146 (54.5) | 2,897 (58.4) |
| High: Degree level | 1,186 (22.7) | 57 (21.3) | 1,129 (22.8) |

Distributions of offspring, maternal and paternal characteristics of the study population.
Abbreviations: N = Number, SD = Standard deviation, MIH = Molar-incisor hypomineralisation, BMI = Body mass index, CSE = Certificate of secondary education, Trimester 1 = First 3 months of pregnancy, Trimester 2 = 18 weeks' gestation, Trimester 3 = 32 weeks' gestation.

### BMI and offspring MIH

In the confounder and other parent BMI-adjusted model (model 3), there was a 4% increased odds of offspring MIH for every unit (1kg/m^2^) increase in maternal BMI (OR: 1.04, 95% CI: 1.00-1.08) (**Table 2 and Figure 3A**). In comparison, point estimates for paternal BMI were in the opposite direction (OR: 0.94, 95% CI: 0.89, 1.00). Negative control outcome analyses for maternal BMI and offspring dental trauma showed no difference in odds, with point estimates around the null (OR: 0.99, 95% CI: 0.96, 1.02) (**Table S2 and Figure 3A)**. Results of complete case analyses were broadly consistent (**Table S4**). Taken together, these results provide some evidence of an association between mean maternal BMI and offspring MIH with negative control analyses suggesting that the results are unlikely to be being driven by unmeasured confounders. In analyses of BMI quintiles, we compared linear and categorical models. Whilst there was statistical evidence for a linear trend for maternal BMI (p-value for per fifth increase = 0.04), **Figure 3B and Table S3** shows that this appeared to be driven by the highest quintile (OR for women in the highest BMI quintile versus the lowest: 1.61, 95% CI: 1.02, 2.60). In comparison, there was no evidence of a positive association for any of the paternal BMI quintiles and offspring MIH (**Figure 3B**).

**Table 2:** Negative control exposure analyses.

|  | Model 1<br>OR (95% CI) | Model 2<br>OR (95% CI) | Model 3<br>OR (95% CI) |
| --- | --- | --- | --- |
| **BMI** |  |  |  |
| **Maternal BMI** | **1.03 (1.00-1.06)** | **1.04 (1.00-1.07)** | **1.04 (1.00-1.08)** |
| *N Total* | 4830 | 3582 | 2698 |
| *N Case* | 252 | 184 | 134 |
| **Paternal BMI** | **0.97 (0.92-1.01)** | **0.96 (0.91-1.01)** | **0.94 (0.89-1.00)** |
| *N Total* | 3993 | 2968 | 2713 |
| *N Case* | 202 | 142 | 128 |
| **Smoking** |  |  |  |
| **Maternal Smoking** | **1.41 (1.06, 1.86)** | **0.98 (0.68, 1.38)** | **0.76 (0.46, 1.22)** |
| *N Total* | 5017 | 4025 | 3184 |
| *N Case* | 267 | 210 | 151 |
| **Paternal Smoking** | **1.11 (0.82, 1.48)** | **1.16 (0.81, 1.64)** | **1.21 (0.82, 1.77)** |
| *N Total* | 4250 | 3270 | 3061 |
| *N Case* | 210 | 155 | 144 |
| **Alcohol** |  |  |  |
| **Maternal Alcohol** | **0.87 (0.65-1.17)** | **0.87 (0.64-1.18)** | **0.79 (0.56-1.12)** |
| *N Total* | 4384 | 4025 | 3288 |
| *N Case* | 234 | 210 | 155 |
| **Paternal Alcohol** | **0.83 (0.45-1.70)** | **1.03 (0.48-2.67)** | **1.04 (0.41-3.47)** |
| *N Total* | 4391 | 3270 | 2666 |
| *N Case* | 215 | 155 | 127 |

Negative control exposure analyses showing associations between maternal and paternal exposures (pre-pregnancy BMI, pregnancy smoking and alcohol consumption) with offspring MIH. Results of univariable and multivariable logistic regression analyses shown as odds ratio (OR) and 95% Confidence intervals (95% CI). Model 1 = unadjusted, Model 2 = confounder adjusted: age, education, parity (maternal), pregnancy smoking (BMI and alcohol analyses) and alcohol consumption (BMI and smoking analyses), Model 3 = adjusted for the same confounders as model 2 and additionally for the other parent's exposure during/around pregnancy.

**Figure 1:**
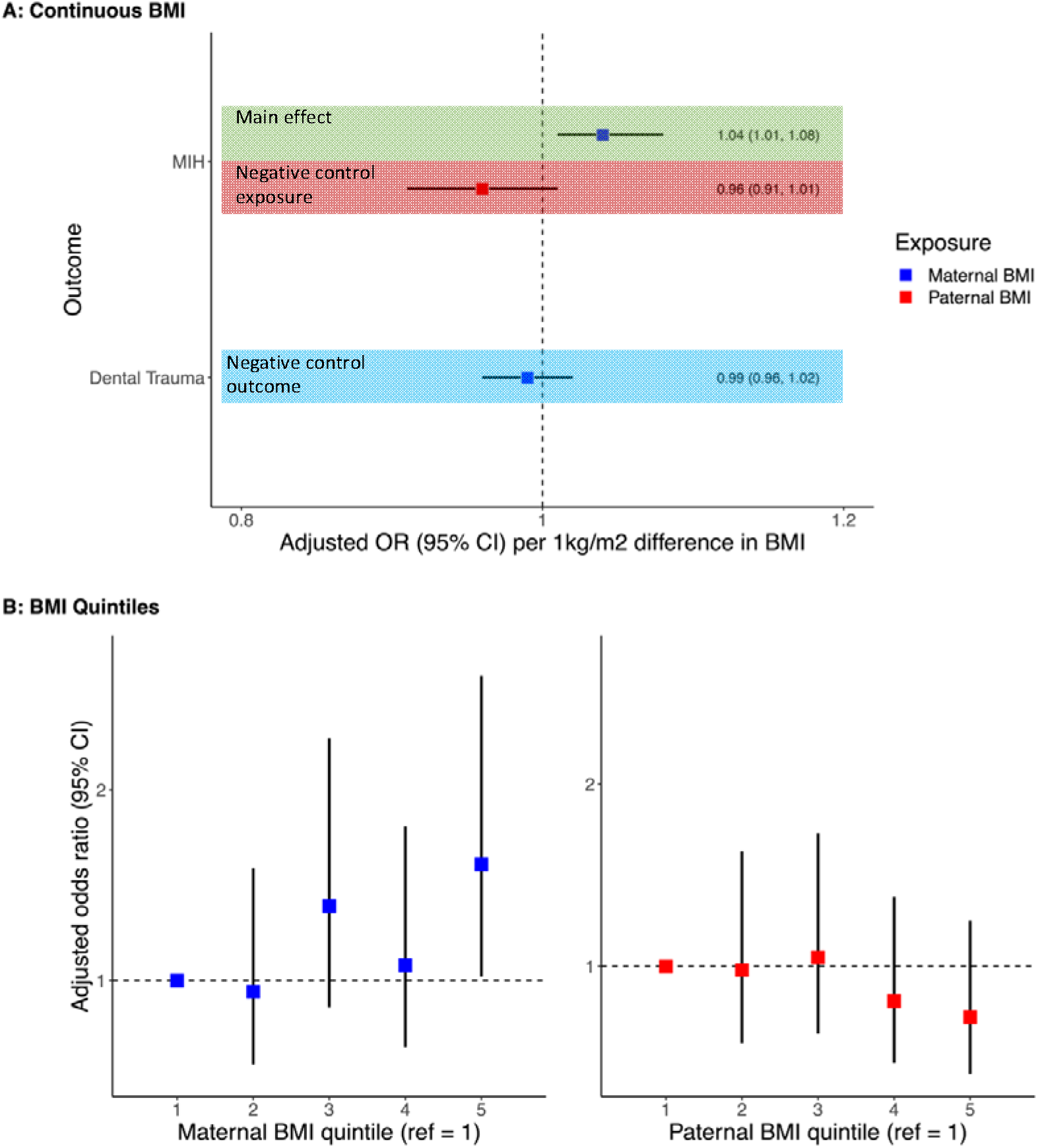
Confounder and other parent body mass index (BMI)-adjusted associations (model 3) for maternal and paternal BMI and offspring molar-incisor hypomineralisation (MIH). Confounders adjusted for include age, parity (maternal), education, pregnancy smoking, pregnancy alcohol consumption and the other parent’s BMI. **A** shows the odds ratio and 95% confidence intervals of offspring MIH for a 1-unit (1kg/m2) difference in maternal BMI (blue) and paternal BMI (red). **B** shows the confounder and other parent BMI-adjusted associations for maternal (red) and paternal BMI (blue) split into quintiles (fifths) and offspring MIH. Results are odds ratio and 95% confidence intervals for BMI quintile and offspring MIH in comparison to BMI quintile 1.

### Smoking and offspring MIH

Unadjusted associations showed maternal pregnancy smoking was associated with an increased odds of offspring MIH (OR: 1.41, 95%CI 1.06, 1.86) (**Table 2)**. However, after adjusting for confounders, the association attenuated to the null (model 2: OR: 0.98, 95% CI: 0.68, 1.38; model 3: OR: 0.76, 95%CI: 0.46, 1.22). Point estimates for paternal smoking were positively associated with MIH, but confidence intervals spanned the null (**model 3**: OR: 1.21, 95%. CI: 0.82, 1.77). For the negative control outcome analyses (**Table S2**), confounder adjusted results similarly showed a small positive association between maternal smoking and offspring dental trauma, and confidence intervals included the null (OR: 1.20, 95%CI: 0.93, 1.53). Taken together, these results provide no evidence of an intrauterine effect of maternal smoking and offspring MIH, although results were imprecisely estimated. In complete case analyses (**Table S3**), all estimates for maternal smoking (including the unadjusted model) were in line with the confounder adjusted main analysis.

### Alcohol consumption and offspring MIH

Associations for alcohol consumption were largely imprecise and showed no clear evidence of an association in maternal (OR: 0.79, 95% CI: 0.56, 1.21) or paternal confounder and other parent adjusted models (OR: 1.04 95% CI: 0.41, 3.47) **(Table 2)**. Similarly, negative control outcome analyses with dental trauma (**Table S2**) showed no clear evidence of an association with maternal pregnancy alcohol consumption (OR: 0.94, 95% CI 0.75, 1.18).

## 4. Discussion

In this prospective cohort study, we found some evidence of a possible intrauterine effect between higher maternal pre-pregnancy BMI and offspring MIH. Additional investigations uncovered that this association appeared to be non-linear and may be driven by a threshold effect in the highest BMI quintile. We did not find similar associations in our negative control analyses, suggesting that these results are unlikely to be explained by confounding. We found no robust evidence to suggest a causal intrauterine effect of maternal pregnancy smoking or alcohol consumption on offspring MIH. Although, we acknowledge that these results were less precise. To our knowledge, this is the first study to use prospective data in large numbers using negative control analyses to explore possible maternal pregnancy risk factors for offspring MIH.

Our results suggested that mothers with a higher BMI may have increased odds of having an offspring with MIH. There are several potential explanations for this; Firstly, FPMs develop *in utero*, therefore it is biologically plausible that maternal pre-pregnancy BMI (which is reflective of weight during early pregnancy) may influence MIH. Pre-pregnancy obesity provides an unfavourable environment for foetal development due to supply of nutrients crossing the placenta in deficit or overabundance (Leddy et al., 2008; Maffeis & Morandi, 2017). This may cause metabolic and physiological changes which alter growth and development (Leddy et al., 2008; Maffeis & Morandi, 2017). A second explanation is that we have not fully accounted for confounders. However, we did attempt to include stringent confounder adjustments and negative control analyses investigations in which our results do not support presence of confounding. Finally, the finding could be a false positive (due to chance). As this is the first study to investigate pre-pregnancy BMI as a potential risk factor for MIH it cannot be compared with previous work. More research with larger studies are warranted to attempt to replicate the association with BMI and to assess the effects of obese and severely obese World Health Organisation categories. If true, our findings would have implications to further support encouragement of a healthy pre-pregnancy BMI in women trying to conceive.

We did not find evidence of an association between maternal pregnancy smoking and offspring MIH. Although unadjusted findings revealed an increased odds of offspring MIH in mothers who smoked, these odds were attenuated close to null after adjusting for confounders. However, these associations were imprecise, therefore we cannot simply reject the possibility of an association between pregnancy smoking and offspring MIH. As these results had limited statistical power, negative control explorations were less meaningful. Our findings support findings of systematic reviews, that there is little evidence for an association between pregnancy smoking on offspring MIH (Fatturi et al., 2019; Silva et al., 2016). A recent well-powered case-control study, not included in these reviews, found evidence of an association between pregnancy smoking and offspring MIH (Lee et al., 2020); however, their exposure data was collected retrospectively, and is therefore susceptible to information bias (Sutton-Tyrrell, 1991). They also did not account for the range of confounders we have included here (e.g. parity and alcohol consumption) which in our study contributed to the full attenuation of the smoking effect on MIH.

Point estimates for maternal alcohol consumption were in the protective direction on offspring MIH, but these findings were imprecise. Therefore, no clear evidence of an association was found, and negative control explorations were not required to explore residual confounding. A protective association seems unlikely as there is no plausible biological explanation. Results from other studies are conflicting and lack statistical power (Fatturi et al., 2019), therefore the effects of alcohol consumption on MIH remains unclear.

A key strength of this study is the use of prospective cohort data which limits recall bias, compared to most studies in this field which have used retrospective study designs, in which recall bias may influence results. Other strengths of this study include the adjustment of a wide range of prospectively measured, relevant confounders, and the use of negative control analyses to detect any residual confounding, which no previous study has accounted for. Taken together, these steps improved robustness of this study, which strengthens the evidence of our findings.

An important limitation to consider is our definition of MIH, “6-year molars that came through brown” at age 7. This may be oversimplistic in comparison to the gold standard EAPD classification (Weerheijm, 2003), and prone to inaccuracy due to the lack of a clinical diagnosis. This means that our MIH cases may include differentials that affect all teeth, such as amelogenesis imperfecta, fluorosis, enamel hypoplasia and caries. Together, these may result in a misclassification of MIH; however, this will be non-systematic with respect to the exposures and would, at worst, attenuate the magnitude of any association towards the null. We intended to prevent misclassification by conducting a sensitivity analysis, including cases with affected molars and incisors. However, with the sample size being small (40/297, 18.8%), additional analyses would not have been particularly informative. Other limitations include self-reporting of data which may have resulted in underreporting of smoking and alcohol consumption; however, this would be expected to weaken any true effect of these exposures towards the null, rather than create false positives.

Another important consideration is the use of negative control analyses; although negative controls are useful, a limitation of this method is that if an association is found with the presence of unmeasured confounding, we cannot specifically know what the confounding may be (Sanderson et al., 2018). This is because this method only detects the presence of confounding, and is therefore considered a “blunt tool” (Lipsitch et al., 2010).

Overall, this study adds to the limited evidence investigating the effects of modifiable maternal risk factors on offspring MIH. This is beneficial, as previous research primarily focuses on non-modifiable risk factors such as maternal and childhood illnesses (Fatturi et al., 2019; Silva et al., 2016), for which intervention is difficult. As the aetiology of MIH is likely to be multifactorial, future research exploring modifiable maternal risk factors, such as BMI, smoking and alcohol consumption, may enable preventative strategies or remove stigma from mothers of children with MIH. To improve comparability and accuracy, it is advisable for future studies to use prospectively obtained data with larger samples, clinically diagnose MIH using the EAPD criteria (Weerheijm, 2003), adjust for relevant confounding variables, and use epidemiological methods to improve causality (eg. negative control analyses) as this present study has. As large enough sample sizes may be difficult to obtain, cross-cohort studies may be an alternative approach to gain sufficient statistical power.

## 5. Conclusion

In conclusion, the findings presented in this study showed some evidence of an intrauterine effect of higher maternal BMI on offspring MIH. We did not find robust evidence for an effect of maternal pregnancy smoking or alcohol consumption on risk of offspring MIH. Further studies with larger numbers utilising prospectively collected data with advanced epidemiological methods and formal MIH diagnoses are required to replicate and further investigate these findings. Furthermore, exploring possible mechanisms that link the pregnancy environment to offspring MIH could identify targets for interventions for its prevention.

## Data Availability

The data that support the findings form this study are available from the ALSPAC cohort (http://www.bristol.ac.uk/alspac/) but restrictions apply to the availability of these data, which were used under license for the current study, and so are not publicly available. Data are however available upon reasonable request and with permission of ALSPAC executive. Please note that the study website contains details of all the data that is available through a fully searchable data dictionary and variable search tool (http://www.bristol.ac.uk/alspac/researchers/our-data/).

http://www.bristol.ac.uk/alspac/researchers/our-data/

http://www.bristol.ac.uk/alspac/

## Acknowledgements

We are extremely grateful to all the families who took part in this study, the midwives for their help in recruiting them, and the whole ALSPAC team, which includes interviewers, computer and laboratory technicians, clerical workers, research scientists, volunteers, managers, receptionists and nurses.

## Abbreviations

ALSPAC: Avon longitudinal study of adults and children
BMI: Body mass index
CI: Confidence interval
FPM: First permanent molars
MIH: Molar-incisor hypomineralisation
OR: Odds ratio

