## Supplementary Material for "The effects of modifiable maternal pregnancy exposures on offspring molar-incisor hypomineralisation: A negative control study"

### **ALSPAC Questionnaires**

#### **Figure S1: “My Teeth” questionnaire completed at age 7**

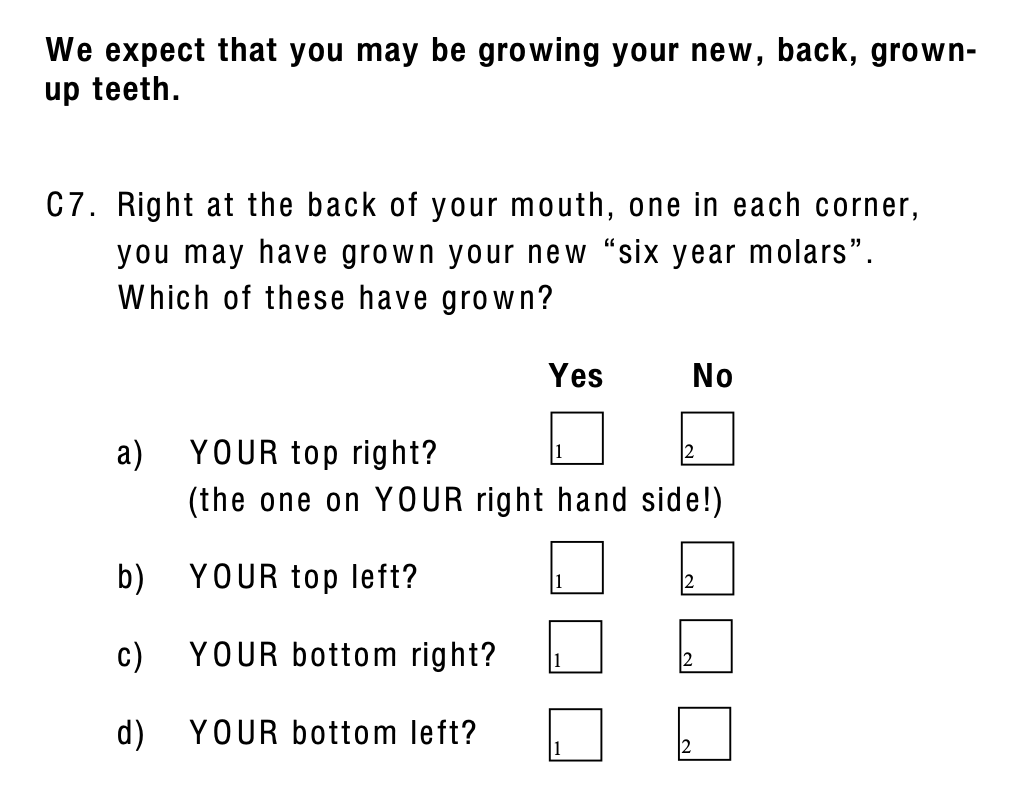

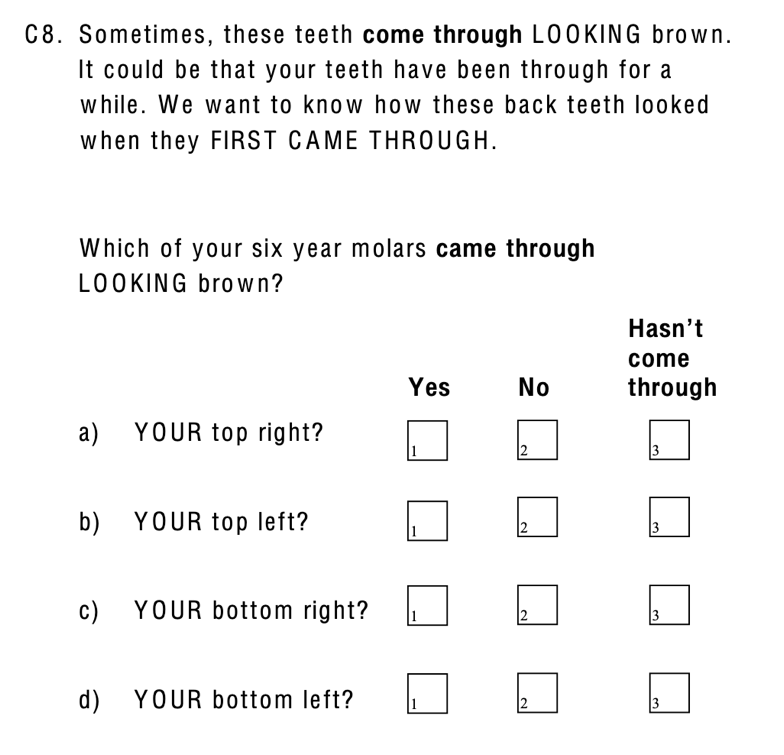

**S1B:**

**S1A:**

**Figure S1: S1A** shows questions used for participant inclusion. **S1B** shows questions used to define cases of MIH, “any child who had at least one molar tooth which came through looking brown”.

#### **Figure S2: “Teeth and Things” questionnaire completed at age 10**

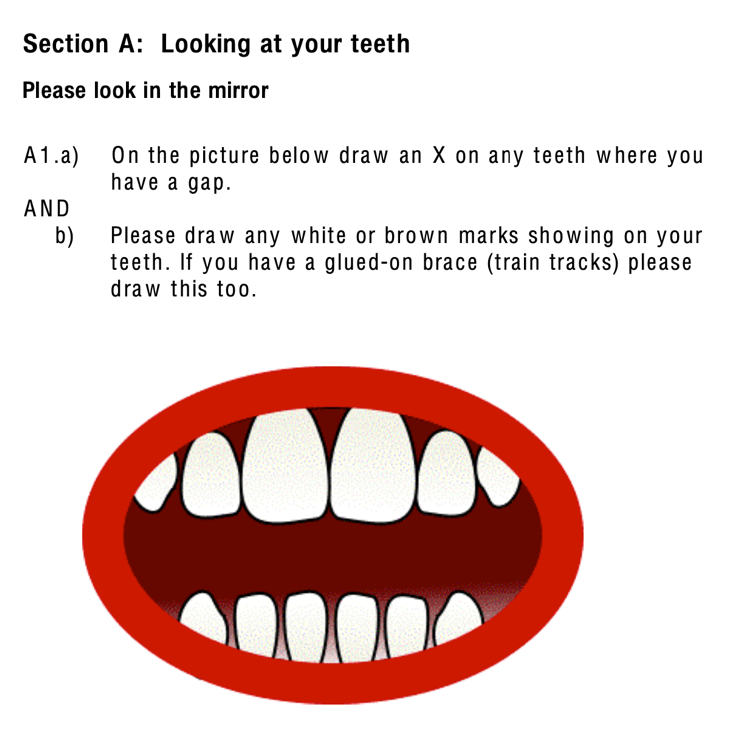

**S2A:**

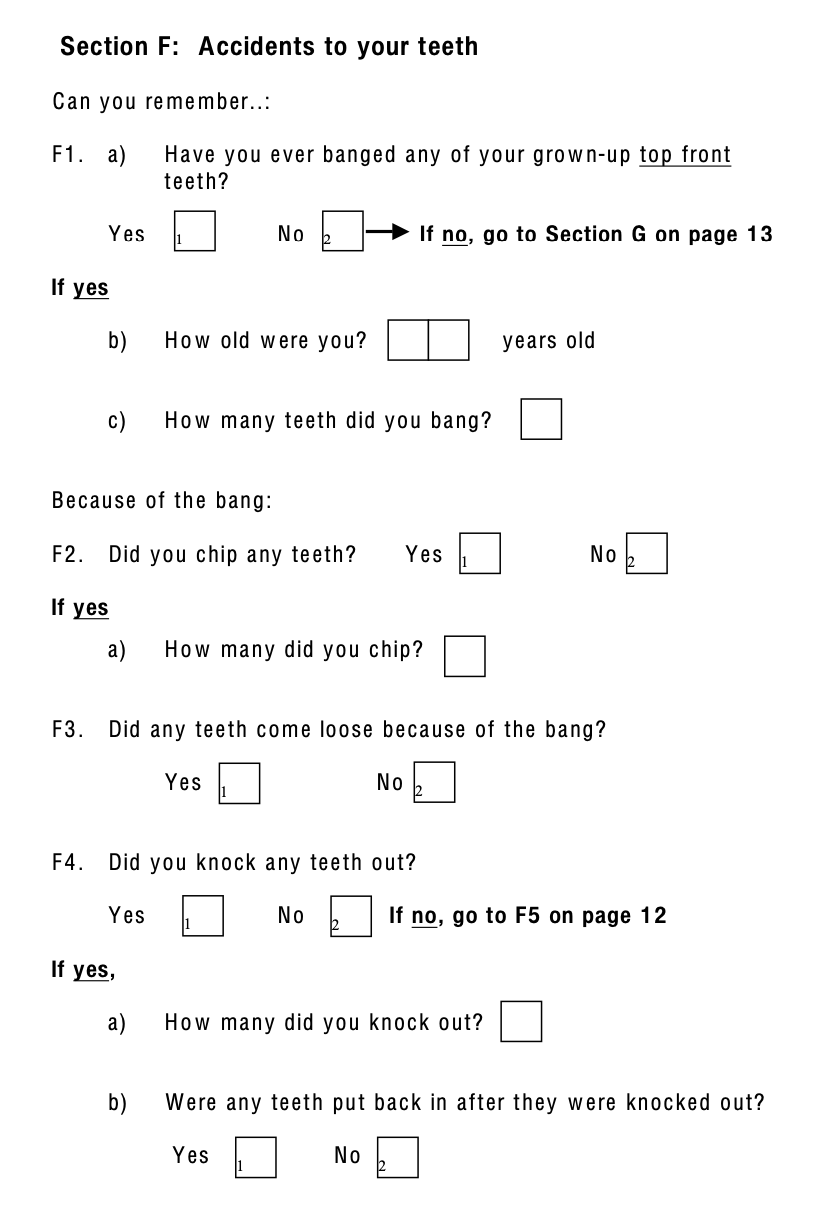

**S2B:**

**Figure S2: S2A shows** questions used for MIH sensitivity analysis, “any child with brown molars and incisors”. **S2B: shows** questions used to define offspring dental trauma, “any child who banged their top adult teeth, which became loose, chipped or knocked out”

| Table S1: Missing data | |
| --- | --- |
| Variable | **N missing (N = 5536 total)** |
| *Offspring* | |
| MIH case | 0 |
| MIH sensitivity | 84 |
| Dental trauma | 922 |
| *Maternal* |  |
| Age | 0 |
| BMI | 706 |
| Pregnancy Smoking |  |
| Trimester 1 | 119 |
| Trimester 2 | 119 |
| Trimester 3 | 590 |
| Any smoking | 519 |
| Pregnancy Alcohol | |
| Trimester 1 | 138 |
| Trimester 2 | 5536 |
| Trimester 3 | 2469 |
| Any alcohol | 1152 |
| Parity | 178 |
| Education | 185 |
| *Paternal* | |
| Age | 1734 |
| BMI | 1543 |
| Pregnancy Smoking | 1286 |
| Pregnancy Alcohol | 1145 |
| Education | 311 |
| Summary of offspring, maternal and paternal missing data obtained from the ALSPAC cohort. N = number, MIH = Molar-incisor hypomineralisation, BMI = Body mass index, Trimester 1 = First 3 months of pregnancy, Trimester 2 = 18 weeks’ gestation, Trimester 3 = 32 weeks’ gestation. | |

| Table S2: Negative control outcome analyses | | |
| --- | --- | --- |
|  | **Model 1**  OR (95% CI) | **Model 2**  OR (95% CI) |
| BMI |  |  |
| Offspring MIH | **1.03 (1.00-1.06)** | **1.04 (1.00-1.07)** |
| *N Total* | 4830 | 3582 |
| *N Case* | 252 | 184 |
| Offspring Dental Trauma | **1.00 (0.98-1.03)** | **0.99 (0.96-1.02)** |
| *N Total* | 4048 | 3037 |
| *N Case* | 537 | 412 |
| Smoking |  |  |
| Offspring MIH | **1.41 (1.06, 1.86)** | **0.98 (0.68, 1.38)** |
| *N Total* | 5017 | 4025 |
| *N Case* | 267 | 210 |
| Offspring Dental Trauma | **1.28 (1.03, 1.50)** | **1.20 (0.93, 1.53)** |
| *N Total* | 4203 | 3402 |
| *N Case* | 566 | 468 |
| Alcohol |  |  |
| Offspring MIH | **0.87 (0.65-1.17)** | **0.87 (0.64-1.18)** |
| *N Total* | 4384 | 4025 |
| *N Case* | 234 | 210 |
| Offspring Dental Trauma | **0.96 (0.78-1.20)** | **0.94 (0.76-1.18)** |
| *N Total* | 3695 | 3402 |
| *N Case* | 505 | 468 |
| Negative control outcome analyses showing associations between maternal BMI and offspring MIH, compared with offspring dental trauma. Results of univariable and multivariable logistic regression analyses, presented as odds ratios (OR) and 95% confidence intervals (CI) for offspring outcome. Model 1 = unadjusted, Model 2 = confounder adjusted: age, education, parity (maternal), smoking and/ or alcohol consumption during pregnancy. | | |

| Table S3: BMI quintile analyses | | | | | |
| --- | --- | --- | --- | --- | --- |
| BMI Quintile | **1** | **2** | **3** | **4** | **5** |
|  | **OR** | **OR** | **OR** | **OR** | **OR** |
|  | (95% CI) | (95% CI) | (95% CI) | (95% CI) | (95% CI) |
| Maternal BMI | Ref. | 0.94 | 1.04 | 1.08 | 1.61 |
|  |  | (0.56, 1.59) | (1.00-1.08) | (0.65, 1.81) | (1.02, 2.60) |
| Paternal BMI | Ref. | 0.98 | 1.05 | 0.81 | 0.72 |
|  |  | (0.58, 1.63) | (0.63, 1.73) | (0.47, 1.38) | (0.41, 1.25) |
| BMI quintile analyses showing the confounder and other parent BMI-adjusted associations (Model 3) for maternal and paternal BMI split into quintiles (fifths) and offspring MIH. Confounders adjusted for include age, parity (maternal), education, pregnancy smoking, pregnancy alcohol consumption and other parent’s BMI. Result presented are odds ratio and 95% confidence intervals for BMI quintile and offspring MIH in comparison to BMI quintile 1 (reference). | | | | | |

| **Table S4: Complete case analyses** | | | |
| --- | --- | --- | --- |
|  | **Model 1** | **Model 2** | **Model 3** |
| **BMI** | OR (95% CI) | OR (95% CI) | OR (95% CI) |
| **Maternal** **BMI** | **1.03 (1.00-1.06)** | **1.04 (1.00-1.07)** | **1.04 (1.00-1.08)** |
| *N Total* | 4830 | 3582 | 2698 |
| *N Case* | 252 | 184 | 134 |
| **Complete case** | **1.06 (1.02-1.11)** | **1.06 (1.02-1.11)** | **1.07 (1.02-1.12)** |
| *N Total* | 4830 | 3582 | 2698 |
| *N Case* | 252 | 184 | 134 |
| **Paternal** **BMI** | **0.97 (0.92-1.01)** | **0.96 (0.91-1.01)** | **0.94 (0.89-1.00)** |
| *N Total* | 3993 | 2968 | 2713 |
| *N Case* | 202 | 142 | 128 |
| **Complete Case** | **0.97 (0.91-1.04)** | **0.97 (0.91-1.04)** | **0.96(0.89-1.02)** |
| *N Total* | 3993 | 2968 | 2713 |
| *N Case* | 202 | 142 | 128 |
| **Smoking** |  |  |  |
| **Maternal Smoking** | **1.41 (1.06, 1.86)** | **0.98 (0.68, 1.38)** | **0.76 (0.46, 1.22)** |
| *N Total* | 5017 | 4025 | 3184 |
| *N Case* | 267 | 210 | 151 |
| **Complete case** | **0.85 (0.49, 1.38)** | **0.83 (0.48, 1.38)** | **0.77 (0.43, 1.32)** |
| *N Total* | 2499 | 2499 | 2499 |
| *N Case* | 118 | 118 | 118 |
| **Paternal Smoking** | **1.11 (0.82, 1.48)** | **1.16 (0.81, 1.64)** | **1.21 (0.82, 1.77)** |
| *N Total* | 4250 | 3270 | 3061 |
| *N Case* | 210 | 155 | 144 |
| **Complete Case** | **1.13 (0.75, 1.66)** | **1.07 (0.71, 1.60)** | **1.15 (0.74, 1.76)** |
| *N Total* | 2499 | 2499 | 2499 |
| *N Case* | 118 | 118 | 118 |
| **Alcohol** |  |  |  |
| **Maternal Alcohol** | **0.87 (0.65-1.17)** | **0.87 (0.64-1.18)** | **0.79 (0.56-1.12)** |
| *N Total* | 4384 | 4025 | 3288 |
| *N Case* | 234 | 210 | 155 |
| **Complete Case** | **0.84 (0.57-1.26)** | **0.85 (0.57-1.28)** | **0.85 (0.57-1.29)** |
| *N Total* | 2499 | 2499 | 2499 |
| *N Case* | 118 | 118 | 118 |
| **Paternal Alcohol** | **0.83 (0.45-1.70)** | **1.03 (0.48-2.67)** | **1.04 (0.41-3.47)** |
| *N Total* | 4391 | 3270 | 2666 |
| *N Case* | 215 | 155 | 127 |
| **Complete Case** | **0.88 ( 0.36-2.95)** | **0.93 (0.38-3.10)** | **0.99 (0.40-3.31)** |
| *N Total* | 3993 | 2968 | 2713 |
| *N Case* | 202 | 142 | 128 |
| Complete case analyses showing associations between maternal and paternal exposures (BMI, smoking and alcohol) with offspring MIH prevalence in full sample and cases with no missing data. Results of univariable and multivariable logistic regression analyses, presented in terms of estimated odds ratio (OR) and 95% Confidence intervals (95% CI). **Model 1 =** unadjusted, **Model 2 =** confounder adjusted: age, education, parity (maternal), smoking and/ or alcohol consumption during pregnancy, **Model 3 =** adjusted for confounders and other parent’s exposure during pregnancy. | | | |
